# Pandemic related Health literacy – A Systematic Review of literature in COVID-19, SARS and MERS pandemics

**DOI:** 10.1101/2020.05.07.20094227

**Authors:** Jun Jie Benjamin Seng, Cheng Teng Yeam, Caleb Weihao Huang, Ngiap Chuan Tan, Lian Leng Low

## Abstract

**Background:** Health literacy plays an essential role in one’s ability to acquire and understand critical medical information in the COVID-19 infodemic and other pandemics.

**Purpose:** To summarize the assessment, levels and determinants of pandemic related health literacy and its associated clinical outcomes.

**Data sources:** Medline®, Embase®, PsychINFO®, CINAHL®, arXiv, bioRxiv, medRxiv, and Social Science Research Network. The start date was unrestricted and current as of 22 April 2020.

**Study selection:** Studies which evaluated health literacy related to novel coronavirus disease 2019 (COVID-19), Severe Acute Respiratory Syndrome (SARS) or Middle East Respiratory Syndrome (MERS)

**Data extraction:** Data on the characteristics of study designs, instruments, participants and level of health literacy were collected. Items used in instruments were grouped under the themes of knowledge, attitudes and practices. Determinants of health literacy were grouped into five domains (socio-demographic, medical, psychological/psychiatric, health systems related and others).

**Data synthesis:** Of 2,065 articles screened, 70 articles were included. 21, 17 and 32 studies evaluated health literacy related to COVID-19, SARS and MERS, respectively. The rates of low pandemic health literacy ranged from 4.3 to 57.9% among medical-related populations and 4.0% to 82.5% among non-medical populations. Knowledge about symptoms and transmission of infection; worry about infection and, practices related to mask usage and hand hygiene was most frequently evaluated. Socio-demographic determinants of health literacy were most studied, where higher education level, older age and female gender were associated with better health literacy. No studies evaluated outcomes associated with health literacy.

**Limitations:** Non-English articles were excluded.

**Conclusion:** The level of pandemic related health literacy is sub-optimal. Healthcare administrators need to be aware of health literacy determinants when formulating policies in pandemics.

## Introduction

With the rapid progression of the novel coronavirus disease 2019 (COVID-19) into a pandemic infecting over 2.5 million patients worldwide, the need to gather and synthesize health-related information to make timely behaviour changes among people has become quintessential. (1, 2) This comes in the wake of an “infodemic” with evolving scientific knowledge about infections being generated daily, which has led to reversals in infection prevention recommendations made within a short span of time.(2-4) For example, the use of cloth masks during the early stages of the COVID-19 pandemic was discouraged by the World Health Organisation due to uncertainty about its efficacy.(3) However, its potential use in slowing the spread of COVID-19 has led to subsequent recommendations by the US Centers for Disease Control and Prevention for it to be worn by healthy individuals.(4) The ease of access to information via social and online media platforms has also become a double-edged sword in this pandemic where there has been substantial propagation of misinformation.(6) Faced with the continuous influx of information related to this pandemic, an individual’s level of health literacy exerts a vital role in one’s ability to acquire, discern and understand accurate medical information.

Health literacy is broadly defined as the “level of capacity one has to obtain, process and understand basic health information and services needed to make appropriate health decisions.” (6) Inadequate levels of health literacy have remained a pervasive problem worldwide, despite medical advances in the past decades. A review by Paasche-Orlow et al. involving 85 studies showed 26% of people living in the United States of America (USA) had low general health literacy.(7) Similar findings were found in Europe where 47% of the population were shown to have limited health literacy.(8) In the setting of non-communicable diseases, the association between health literacy with increased healthcare costs, morbidity and mortality is well-established. (9) The equal importance of health literacy in communicable diseases was highlighted in the recent COVID-19 crisis and previous coronavirus pandemics such as the Severe Acute Respiratory Syndrome (SARS) and Middle East Respiratory Syndrome (MERS).(1, 10) In contrast to general health literacy required for the prevention or management of chronic diseases, these pandemics require an individual’s readiness and adaptive ability in developing their pandemic related and critical health literacy quickly. This is critical as the rapid and successful implementation of infectious diseases control measures requires the collective compliance of all individuals. (11, 12)

Varying levels of pandemic related health literacy have been reported. In a study which examined COVID-19 awareness and attitudes among chronic disease patients in the USA, it was worrisome to note that one-third of participants were unable to identify symptoms associated with COVID-19 and 24.6% of participants felt that they were not likely to contract the virus.(13) Another study conducted by Roy et al. showed that only 43% of responders regarded COVID-19 as a contagious disease and 18.2% regarded fever as a symptom of COVID-19. (14) In contrast, a study in China showed that health literacy was high among participants, where a 90% accuracy rate was reported for the COVID-19 knowledge questionnaire administered. For other coronavirus pandemics such as SARS and MERS, differences in pandemic related literacy levels across different study populations were also reported.(15, 16)

Variations in general health literacy have been linked to multiple determinants ranging from education to socioeconomic statuses.(17) Likewise, this is expected for pandemic related health literacy. Understanding the levels and determinants of pandemic related health literacy across different populations is essential for healthcare policymakers to formulate optimal strategies for effective communication of critical medical information in the COVID-19 crisis and future pandemics.

Hence, the objective of this review is to evaluate and summarize the assessment and level of health literacy related to COVID-19, SARS and MERS and its associated determinants. In the absence of a gold standard instrument, the themes identified from items used in the health literacy instruments across studies will guide the development of future pandemic related health literacy instruments. The secondary objective of this study was to evaluate the clinical outcomes associated with poor pandemic related health literacy.

## Methodology

This systematic review has been registered on PROSPERO (Registration number: CRD 42020181171).

### Data sources and searches

The literature search was conducted in Medline®, Embase®, PsychINFO® and CINAHL®, in accordance with the Preferred Reporting Items for Systematic Reviews and Meta-Analyses (PRISMA) checklist. Due to the relative novelty and recent nature of the COVID-19 pandemic, pre-prints from four widely used databases which included arXiv, bioRxiv, medRxiv, and Social Science Research Network (SSRN) were extracted for evaluation. Keywords employed in the search strategy included terms related to health literacy as well as the viruses and syndromes implicated in the three coronavirus pandemics which were namely COVID-19, MERS and SARS. Terms related to health literacy were adapted from reviews which evaluated health literacy in other patient populations. (18, 19) The full search strategy was detailed in Supplementary File 1. The start date of the search was unrestricted and current as of 22 April 2020.

### Study selection

Full-text articles, both peer-reviewed and non-peer reviewed in the English language were retrieved from the eight databases. Studies which evaluated health literacy related to COVID-19, SARS or MERS among adult participants aged ≥18 years old from the general population, healthcare sectors and infected patients were included. For the study designs, both interventional and observational studies such as cohort, cross sectional and case control studies were included. Case series, case reports, other irrelevant meta-analyses and systematic reviews were excluded. We also excluded studies which evaluated paediatric populations and non-human subjects.

Two independent reviewers (JJB Seng and CT Yeam) performed the screening and inclusion of articles. All disagreements encountered during the review process were discussed. In situations where the disagreements could not be resolved, a third independent reviewer (CWH Huang) arbitrated to achieve consensus.

### Data extraction and Quality assessment

Data extracted included the socio-demographic and clinical characteristics of the study participants such as their age, race/ethnicity, education levels, income levels, study designs, instruments used for assessment of health literacy, the definition of health literacy used in studies, level of health literacy, factors associated with health literacy and clinical outcomes associated with health literacy.

For the risk of bias assessment, the Quality Assessment Tool for Studies by National Health, Lung and Blood Institute was adopted to evaluate the methodological quality of included articles.(20) Each study is rated as low, moderate and high risk of bias by the two independent reviewers (JJB Seng and CT Yeam) based on the responses obtained from the ten items. In situations where insufficient information was available to score an item, the authors of the study were contacted for clarification. If the authors could not be contacted, the item was rated as high risk of bias. All disagreements were resolved via discussion between the two reviewers. Only studies which were rated as low and moderate risk of bias were included in this review.

### Data Synthesis and Analysis

Descriptive statistics were used to summarize the characteristics of included studies. With regards to the level of health literacy, we reported the average percentage of correct answers or the percentage of participants with low health literacy as defined by cut-offs described in each study, where available. As there are no gold-standard health literacy instruments developed for COVID-19, SARS or MERS(21), significant heterogeneity is expected in the types of tools used for assessment of health literacy across participants. Consequently, meta-analysis could not be performed. Questions from instruments used across included studies were classified into three main themes, which were 1) knowledge, 2) attitudes and 3) practices, to help guide future development of standardised COVID-19 and pandemic health literacy tools. The analyses were segregated by medical and non-medical populations due to the expected differing levels of health literacy in the two populations. For studies where the questionnaires were not available, study authors were contacted for the questionnaire. If there were no replies from the authors, the themes were extracted from the description of the questionnaires in the main text. A framework for core items to be included in pandemic health literacy tools was also proposed based on common themes assessed across studies.

For factors associated with better health literacy, they were categorized into five domains which encompassed socio-demographic, medical, psychological/psychiatric, health systems related and others. A narrative review was provided for the factors evaluated among included studies. Clinical outcomes associated with poor health literacy among patients infected with COVID-19, MERS and SARS included time from illness onset to seeking medical treatment, hospitalisation and duration of hospitalisation, admission to intensive care units and length of ICU stay, need for ventilator support, recovery from infection and re-infection.

### Funding source

This study was not funded by any organisation.

## Results

Figure 1 shows the flowchart for the inclusion of articles. A total of 1,965 published articles and 40 pre-prints were retrieved. After removal of duplicates, exclusion of irrelevant articles and inclusion of articles identified from hand-searching, a total of 70 articles were included in this review. The percentage of concordance during the initial article screening was 90%. Details pertaining to the study designs and characteristics of participants among included studies were reported in Supplementary File 2. For the risk of bias, 48 (68.5%) and 22 (31.4%) studies were rated as low and moderate risk of bias. No studies were rated as high risk of bias (Supplementary File 3). Table 1 shows a summary of the characteristics of the included studies. Majority of included studies were cross-sectional in design (n=65, 92.9%) and were conducted during the pandemics (n=69, 98.6%). 21 (30%) studies recruited more than 1000 participants.

**Figure 1.**
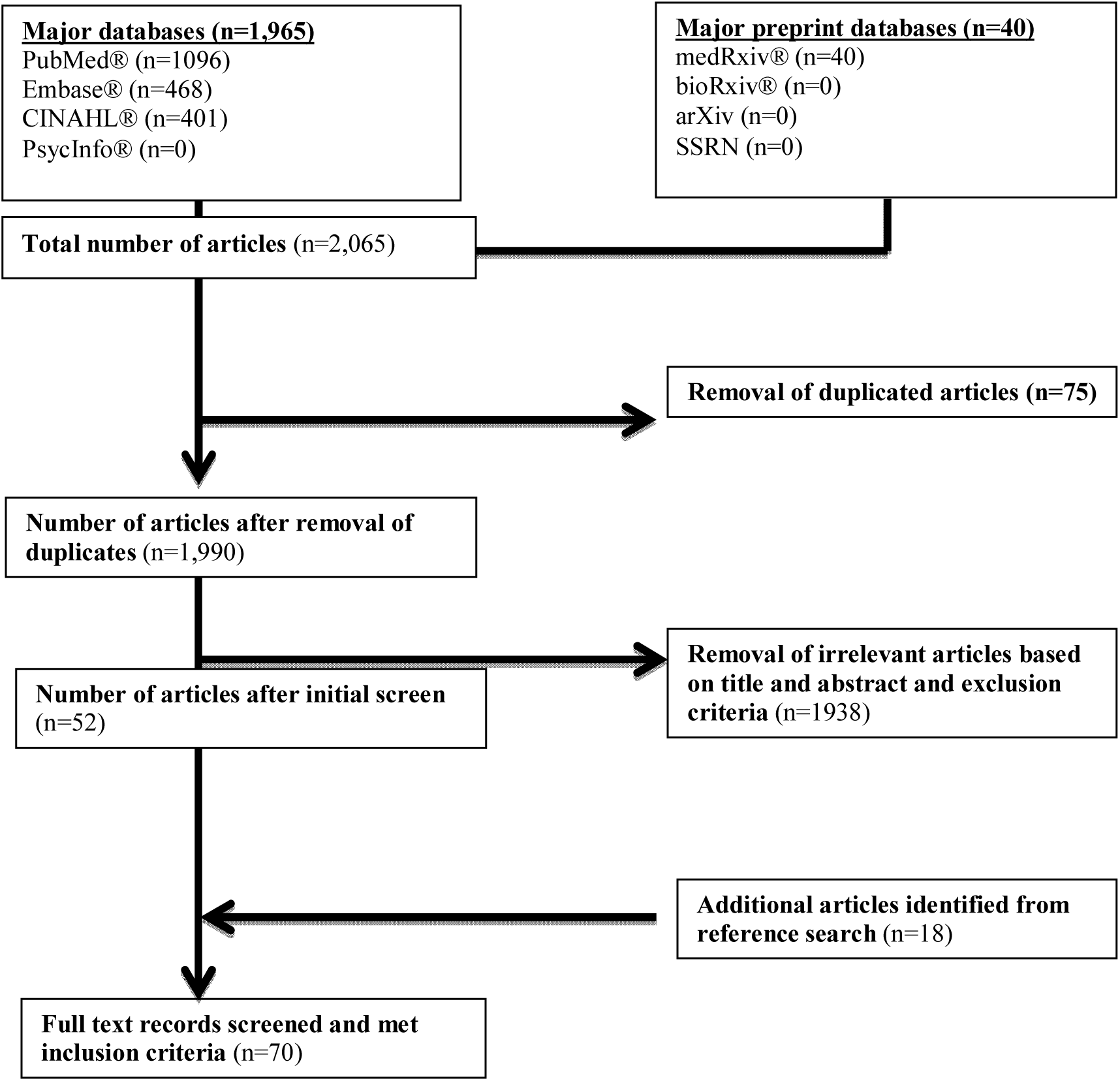
Flow chart for the inclusion of articles for review

**Table 1.** Overview of included studies (n=70)

| Variables | Number of studies, (%) |  |  |  |
| --- | --- | --- | --- | --- |
|  | COVID-19<br>(n=21) | SARS<br>(n=17) | MERS<br>(n=32) | Overall<br>(n=70) |
| **Period of study** |  |  |  |  |
| During epidemic/pandemic | 21 (100) | 16 (94.1) | 32 (100) | 69 (98.6) |
| After epidemic/pandemic | 0 (0) | 1 (5.9) | 0 (0) | 1 (1.4) |
| **Continent of study** <sup>†</sup> |  |  |  |  |
| Asia | 15 (71.4) | 14 (82.4) | 30 (93.8) | 59 (84.3) |
| Europe | 1 (4.8) | 2 (11.8) | 2 (6.3) | 5 (7.1) |
| North America | 3 (14.3) | 1 (5.9) | 0 (0) | 4 (5.7) |
| Multi-continent | 1 (4.8) | 0 (0) | 0 (0) | 1 (1.4) |
| Africa | 1 (4.8) | 0 (0) | 0 (0) | 1 (1.4) |
| **Country of study** |  |  |  |  |
| Saudi Arabia | 0 (0) | 0 (0) | 27 (84.4) | 27 (38.6) |
| Hong Kong | 0 (0) | 7 (41.2) | 0 (0) | 7 (10) |
| India | 4 (19) | 0 (0) | 0 (0) | 4 (5.7) |
| USA | 3 (14.3) | 1 (5.9) | 0 (0) | 4 (5.7) |
| Multi-country | 3 (14.3) | 1 (5.9) | 0 (0) | 4 (5.7) |
| Singapore | 0 (0) | 3 (17.6) | 1 (3.1) | 4 (5.7) |
| China | 3 (14.3) | 0 (0) | 0 (0) | 3 (4.3) |
| Iran | 2 (9.5) | 0 (0) | 0 (0) | 2 (2.9) |
| France | 0 (0) | 0 (0) | 2 (6.3) | 2 (2.9) |
| Korea | 0 (0) | 0 (0) | 2 (6.3) | 2 (2.9) |
| Others‡ | 6 (28.6) | 5 (29.4) | 0 (0) | 11 (15.7) |
| **Type of study** |  |  |  |  |
| Cross-sectional studies | 21 (100) | 15 (88.2) | 29 (90.6) | 65 (92.9) |
| Interventional studies | 0 (0) | 1 (5.9) | 3 (9.4) | 4 (5.7) |
| Qualitative studies | 0 (0) | 1 (5.9) | 0 (0) | 1 (1.4) |
| **Sample size** |  |  |  |  |
| 0-499 | 7 (33.3) | 9 (52.9) | 21 (65.6) | 37 (52.9) |
| 500-1,000 | 4 (19) | 3 (17.6) | 4 (12.5) | 11 (15.7) |
| 1,000-5,000 | 8 (38.1) | 4 (23.5) | 7 (21.9) | 19 (27.1) |
| 5,001-10,000 | 2 (9.5) | 1 (5.9) | 0 (0) | 3 (4.3) |
| **Study population** |  |  |  |  |
| *Non-medical personnel* |  |  |  |  |
| • General population | 10 (47.6) | 8 (47.1) | 8 (25) | 26 (37.1) |
| • University Students | 0 (0) | 0 (0) | 4 (12.5) | 4 (4.3) |
| • Elderly | 0 (0) | 3 (17.6) | 0 (0) | 3 (2.9) |
| • Rural villagers | 0 (0) | 1 (5.9) | 0 (0) | 1 (1.4) |
| • Patients (Dental) | 0 (0) | 1 (5.9) | 1 (3.1) | 2 (2.9) |
| • Pilgrims | 0 (0) | 0 (0) | 1 (3.1) | 1 (1.4) |
| *Medical personnel* |  |  |  |  |
| • Healthcare professionals | 9 (42.9) | 3 (17.6) | 11 (34.4) | 23 (32.9) |
| • Medicine students | 2 (9.5) | 0 (0) | 6 (18.8) | 8 (11.4) |
| Mixed study populations§ | 0 (0) | 1 (5.9) | 1 (3.1) | 1 (1.4) |
| **Rate of response in studies** |  |  |  |  |
| Not specified | 10 (47.6) | 6 (35.3) | 11 (34.4) | 27 (38.6) |
| 0-25% | 2 (9.5) | 0 (0) | 1 (3.1) | 4 (5.7) |
| 25.1 – 50% | 0 (0) | 1 (5.9) | 4 (12.5) | 5 (7.1) |
| 50.1% - 75% | 3 (14.3) | 6 (35.3) | 4 (12.5) | 12 (17.1) |
| 75-100% | 6 (28.6) | 4 (23.5) | 12 (37.5) | 22 (31.4) |
| **Modality of assessment** |  |  |  |  |
| Questionnaires |  |  |  |  |
| - Online questionnaires | 20 (95.2) | 2 (11.8) | 5 (15.6) | 27 (38.6) |
| - Physical questionnaires | 0 (0) | 3 (17.6) | 21 (65.6) | 24 (34.3) |
| Interviews |  |  |  |  |
| - Face to face | 0 (0) | 6 (35.3) | 6 (18.8) | 12 (17.1) |
| - Telephone | 1 (4.8) | 6 (35.3) | 0 (0) | 7 (10) |
| **Language of Questionnaire** |  |  |  |  |
| English | 16 (76.2) | 8 (47.1) | 9 (28.1) | 33 (47.1) |
| Non-English | 5 (23.8) | 8 (47.1) | 15 (46.9) | 28 (40) |
| Multiple languages | 0 (0) | 1 (5.9) | 8 (25) | 9 (12.9) |
| **Validated Instruments** |  |  |  |  |
| Yes | 7 (33.3) | 4 (23.5) | 11 (34.4) | 22 (31.4) |
| No | 14 (66.7) | 13 (76.5) | 21 (65.6) | 48 (68.6) |
| **Number of health literacy questions** |  |  |  |  |
| Not available | 0 (0) | 3 (17.6) | 0 (0) | 3 (4.3) |
| 1-10 | 3 (14.3) | 6 (35.3) | 3 (9.4) | 12 (17.1) |
| 11-20 | 8 (38.1) | 3 (17.6) | 19 (59.4) | 30 (42.9) |
| 21-30 | 7 (33.3) | 1 (5.9) | 6 (18.8) | 14 (20) |
| ≥31 | 3 (14.3) | 4 (23.5) | 4 (12.5) | 11 (15.7) |
| **Components of health literacy addressed** |  |  |  |  |
| Knowledge | 20 (95.2) | 14 (82.4) | 31 (96.9) | 65 (92.9) |
| Attitudes | 17 (81) | 11 (64.7) | 21 (65.6) | 49 (70) |
| Behaviours / Practices | 14 (66.7) | 6 (35.3) | 11 (34.4) | 31 (44.3) |

<sup>†</sup> There was no study from Australia and South America.
<sup>‡</sup> Other countries included Qatar, Pakistan, UAE, Vietnam, Taiwan, Japan, Nigeria, Malaysia, Netherlands, Italy and Jordan (n=1 for listed countries).
<sup>§</sup> A study evaluated both the elderly population and healthcare professionals
<sup>||</sup> Language combinations included English and Arabic; Japanese and English; Chinese, English and Malay
**Abbreviations:** COVID-19 - novel coronavirus 2019; SARS – Severe Acute Respiratory Syndrome; MERS – Middle East Respiratory Syndrome

### COVID-19

A total of 21 (30.0%) studies examined health literacy related to COVID-19 (Table 1). Majority of the studies were conducted in Asia (71.4%) and North America (14.3%). Most studies were conducted among the general population (n=10, 47.6%). The primary mode of health literacy assessment across studies was via online questionnaires (n=20, 95.2%). Aspects of health literacy that were assessed in the instruments included knowledge (n=20, 95.2%), attitudes (n=17, 81%) and practices (n=14, 66.7%), of which only 7 (33.3%) studies performed validation of their questionnaire. Most questionnaires (n=8, 38.1%) contained 11-20 items. Pertaining to health literacy, the average percentage of correct answers among medical personnel ranged from 67.0 to 94.8%, and low health literacy was reported among 5.8 to 43.5% of participants (Supplementary File 2). For non-medical populations, their scores ranged from 62.9 to 90.0%, and the proportion of participants with low health literacy was estimated at 16.1% (Supplementary File 2).

### SARS

Seventeen (24.2%) studies evaluated the level of health literacy related to SARS (Table 1). Majority of studies were mostly performed in Asia (82.4%), Europe (11.8%) and North America (5.9%). The most common groups of study participants included the general population (n=8, 58.8%) and healthcare professionals (n=3, 17.6%). For the assessment of health literacy, these were conducted primarily via interviews (n=12, 70.6%) and questionnaires (n=5, 29.4%). Aspects of health literacy that were evaluated in the instruments were knowledge (n=14, 94.1%), attitudes (n=11, 64.7%) and behaviour / practices (n=6, 82.4). Majority of the instruments were not validated (n=13, 76.5%). Among the number of health literacy questions, the majority (n=6, 35.3%) had 1-10 questions, while four studies employed ≥31 questions. Pertaining to health literacy, the average correct answers from medical personnel ranged from 53.0 to 70.4%, and participants with low health literacy were estimated at 28% (Supplementary File 2). For non-medical personnel, the average correct answers ranged from 42.3 to 93.1%, and participants with low health literacy ranged from 39.9 to 82.5% (Supplementary File 2).

### MERS

Among the pandemics, the most number of studies examined health literacy related to MERS (n=32, 45.7%) (Table 1). The studies were mostly conducted in Asia (n=30, 93.8%) and Europe (n=2, 6.3%) where the majority of them were conducted in Saudi Arabia (n=27, 84.4%). The most common group of participants recruited comprised of healthcare professionals (n=11, 34.4%) and medical students (n=6, 18.8%). The assessment of health literacy was conducted via physical questionnaires (n=21, 37.5%) and face to face interviews (n=6, 18.8%) predominantly. Aspects of health literacy that was most frequently evaluated were knowledge (n=31, 96.9%), attitudes (n=21, 65.6%) and behaviour / practices (n=11, 34.4%). Only 34.4% of the instruments were validated. Pertaining to number of questions in the instruments, most studies utilized 11-20 questions (n=19, 59.4%) and 21-30 questions (n=6, 18.8%). With regards to health literacy, the average correct answers from medical personnel ranged from 42.7 to 96.4%, and participants with low health literacy ranged from 4.3 to 57.9% (Supplementary File 2). For non-medical personnel, the average correct answers ranged from 26.1 to 90.1%, and participants with low health literacy ranged from 4.0 to 59.2% (Supplementary File 2).

### Themes identified from items used in health literacy questionnaires

Among the three themes, pandemic related knowledge was most studied, followed by practices and attitudes (Tables 2-4). In the knowledge domain, symptoms (13, 14, 16, 22-69), transmission (14, 16, 22, 23, 25-29, 31-34, 37-49, 51, 52, 55-57, 59-78) and incubation period of the virus (16, 23, 26-28, 32, 37, 38, 41, 42, 46-50, 52-57, 59-64, 66-69, 78); management and treatment options (14, 16, 22-24, 26, 27, 29, 37, 39-43, 46, 48-51, 55, 56, 58, 60, 62, 64, 66-70, 78, 79); and clinical outcomes associated with infection (13, 16, 23-27, 29, 34, 37, 38, 40, 48, 50-52, 55, 56, 59, 61, 63-67, 69, 74-76, 78-81); high risk populations for infection (16, 23, 26, 27, 29, 37, 39-41, 48, 49, 55-59, 63, 65, 67, 68, 72, 78, 79); availability of vaccine (16, 29, 37, 39, 40, 42-44, 48, 52, 55, 56, 58, 60-63, 65, 68, 70, 74-76, 79); role of hand hygiene(14, 16, 24, 25, 28, 29, 42, 43, 45, 47, 48, 50, 52, 55-58, 60, 64, 66, 69-71, 82, 83) was most studied for medical and non-medical staff. (Table 2) For medical related populations specifically, knowledge about epidemiology (29, 37, 39, 42, 44, 53, 58, 59, 61-63, 65, 66, 78, 79) and diagnosis of infection (16, 46, 48, 49, 55, 56, 58-63, 67, 76) were also frequently evaluated.

**Table 2.**
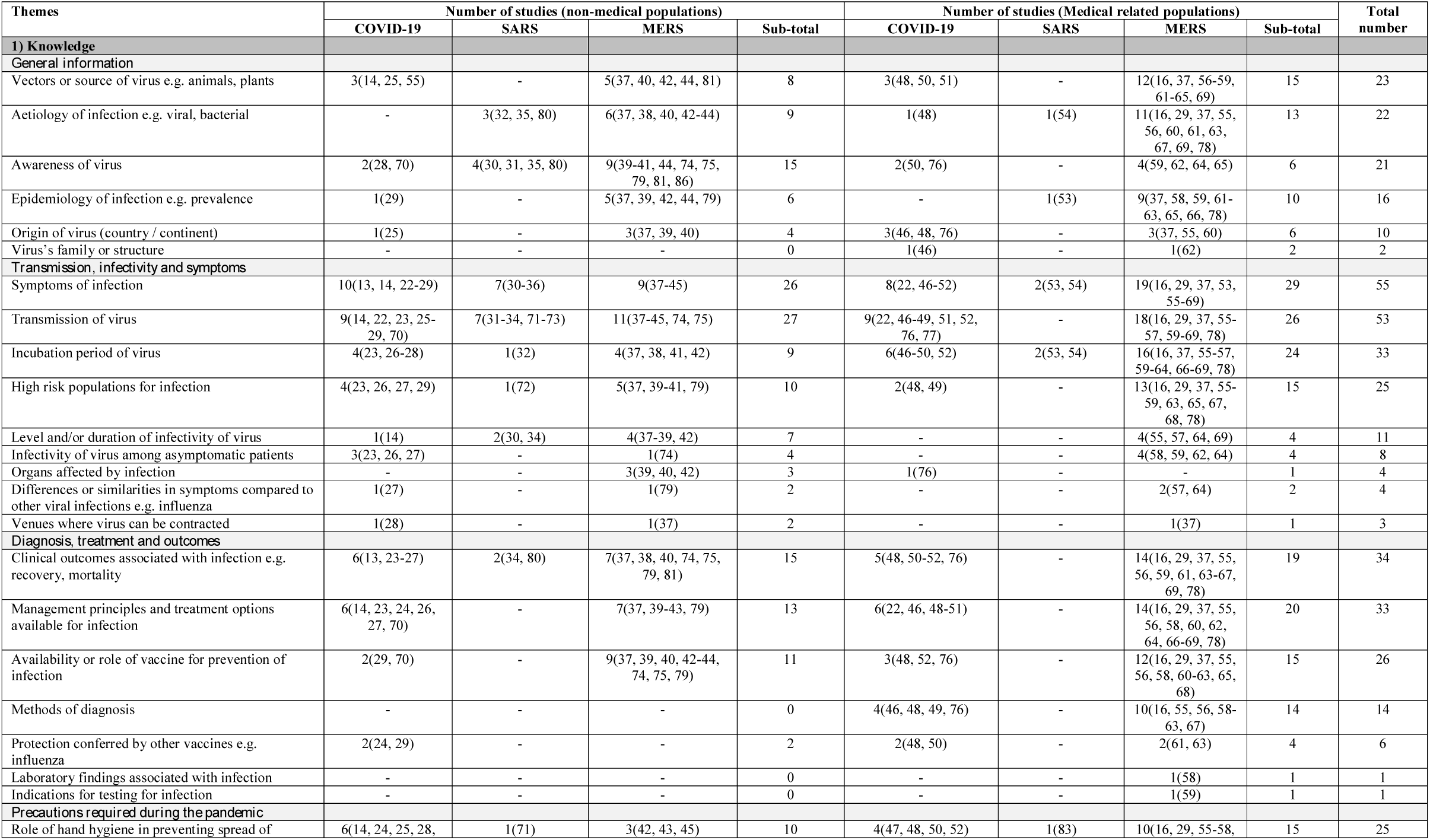
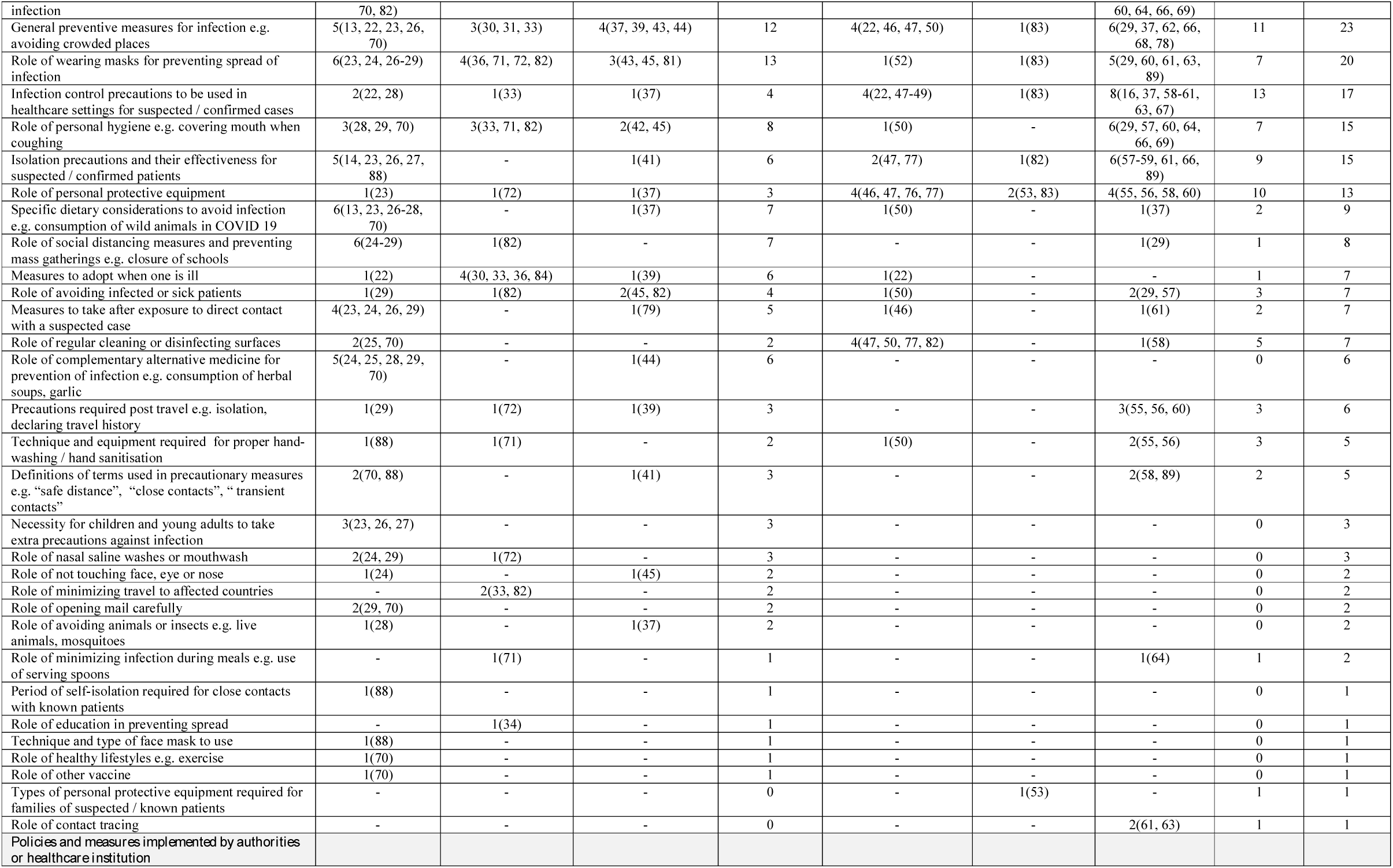
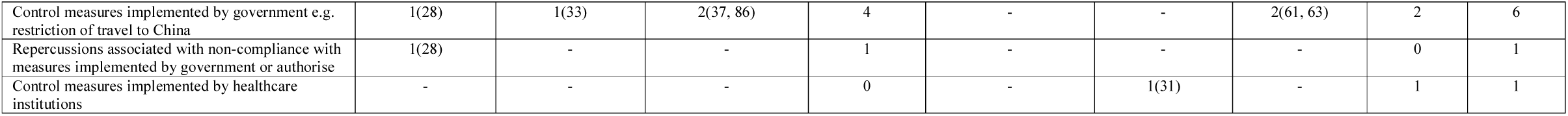
Themes identified from knowledge-based questions from instruments

| Themes | Number of studies (non-medical populations) |  |  |  | Number of studies (Medical related populations) |  |  |  | Total number |
| --- | --- | --- | --- | --- | --- | --- | --- | --- | --- |
|  | COVID-19 | SARS | MERS | Sub-total | COVID-19 | SARS | MERS | Sub-total |  |
| **1) Knowledge** |  |  |  |  |  |  |  |  |  |
| *General information* |  |  |  |  |  |  |  |  |  |
| Vectors or source of virus e.g. animals, plants | 3(14, 25, 55) | - | 5(37, 40, 42, 44, 81) | 8 | 3(48, 50, 51) | - | 12(16, 37, 56-59, 61-65, 69) | 15 | 23 |
| Aetiology of infection e.g. viral, bacterial | - | 3(32, 35, 80) | 6(37, 38, 40, 42-44) | 9 | 1(48) | 1(54) | 11(16, 29, 37, 55, 56, 60, 61, 63, 67, 69, 78) | 13 | 22 |
| Awareness of virus | 2(28, 70) | 4(30, 31, 35, 80) | 9(39-41, 44, 74, 75, 79, 81, 86) | 15 | 2(50, 76) | - | 4(59, 62, 64, 65) | 6 | 21 |
| Epidemiology of infection e.g. prevalence | 1(29) | - | 5(37, 39, 42, 44, 79) | 6 | - | 1(53) | 9(37, 58, 59, 61-63, 65, 66, 78) | 10 | 16 |
| Origin of virus (country / continent) | 1(25) | - | 3(37, 39, 40) | 4 | 3(46, 48, 76) | - | 3(37, 55, 60) | 6 | 10 |
| Virus's family or structure | - | - | - | 0 | 1(46) | - | 1(62) | 2 | 2 |
| *Transmission, infectivity and symptoms* |  |  |  |  |  |  |  |  |  |
| Symptoms of infection | 10(13, 14, 22-29) | 7(30-36) | 9(37-45) | 26 | 8(22, 46-52) | 2(53, 54) | 19(16, 29, 37, 53, 55-69) | 29 | 55 |
| Transmission of virus | 9(14, 22, 23, 25-29, 70) | 7(31-34, 71-73) | 11(37-45, 74, 75) | 27 | 9(22, 46-49, 51, 52, 76, 77) | - | 18(16, 29, 37, 55-57, 59-69, 78) | 26 | 53 |
| Incubation period of virus | 4(23, 26-28) | 1(32) | 4(37, 38, 41, 42) | 9 | 6(46-50, 52) | 2(53, 54) | 16(16, 37, 55-57, 59-64, 66-69, 78) | 24 | 33 |
| High risk populations for infection | 4(23, 26, 27, 29) | 1(72) | 5(37, 39-41, 79) | 10 | 2(48, 49) | - | 13(16, 29, 37, 55-59, 63, 65, 67, 68, 78) | 15 | 25 |
| Level and/or duration of infectivity of virus | 1(14) | 2(30, 34) | 4(37-39, 42) | 7 | - | - | 4(55, 57, 64, 69) | 4 | 11 |
| Infectivity of virus among asymptomatic patients | 3(23, 26, 27) | - | 1(74) | 4 | - | - | 4(58, 59, 62, 64) | 4 | 8 |
| Organs affected by infection | - | - | 3(39, 40, 42) | 3 | 1(76) | - | - | 1 | 4 |
| Differences or similarities in symptoms compared to other viral infections e.g. influenza | 1(27) | - | 1(79) | 2 | - | - | 2(57, 64) | 2 | 4 |
| Venues where virus can be contracted | 1(28) | - | 1(37) | 2 | - | - | 1(37) | 1 | 3 |
| *Diagnosis, treatment and outcomes* |  |  |  |  |  |  |  |  |  |
| Clinical outcomes associated with infection e.g. recovery, mortality | 6(13, 23-27) | 2(34, 80) | 7(37, 38, 40, 74, 75, 79, 81) | 15 | 5(48, 50-52, 76) | - | 14(16, 29, 37, 55, 56, 59, 61, 63-67, 69, 78) | 19 | 34 |
| Management principles and treatment options available for infection | 6(14, 23, 24, 26, 27, 70) | - | 7(37, 39-43, 79) | 13 | 6(22, 46, 48-51) | - | 14(16, 29, 37, 55, 56, 58, 60, 62, 64, 66-69, 78) | 20 | 33 |
| Availability or role of vaccine for prevention of infection | 2(29, 70) | - | 9(37, 39, 40, 42-44, 74, 75, 79) | 11 | 3(48, 52, 76) | - | 12(16, 29, 37, 55, 56, 58, 60-63, 65, 68) | 15 | 26 |
| Methods of diagnosis | - | - | - | 0 | 4(46, 48, 49, 76) | - | 10(16, 55, 56, 58-63, 67) | 14 | 14 |
| Protection conferred by other vaccines e.g. influenza | 2(24, 29) | - | - | 2 | 2(48, 50) | - | 2(61, 63) | 4 | 6 |
| Laboratory findings associated with infection | - | - | - | 0 | - | - | 1(58) | 1 | 1 |
| Indications for testing for infection | - | - | - | 0 | - | - | 1(59) | 1 | 1 |
| *Precautions required during the pandemic* |  |  |  |  |  |  |  |  |  |
| Role of hand hygiene in preventing spread of | 6(14, 24, 25, 28, | 1(71) | 3(42, 43, 45) | 10 | 4(47, 48, 50, 52) | 1(83) | 10(16, 29, 55-58, | 15 | 25 |
| infection | 70, 82) |  |  |  |  |  | 60, 64, 66, 69) |  |  |
| General preventive measures for infection e.g. avoiding crowded places | 5(13, 22, 23, 26, 70) | 3(30, 31, 33) | 4(37, 39, 43, 44) | 12 | 4(22, 46, 47, 50) | 1(83) | 6(29, 37, 62, 66, 68, 78) | 11 | 23 |
| Role of wearing masks for preventing spread of infection | 6(23, 24, 26-29) | 4(36, 71, 72, 82) | 3(43, 45, 81) | 13 | 1(52) | 1(83) | 5(29, 60, 61, 63, 89) | 7 | 20 |
| Infection control precautions to be used in healthcare settings for suspected / confirmed cases | 2(22, 28) | 1(33) | 1(37) | 4 | 4(22, 47-49) | 1(83) | 8(16, 37, 58-61, 63, 67) | 13 | 17 |
| Role of personal hygiene e.g. covering mouth when coughing | 3(28, 29, 70) | 3(33, 71, 82) | 2(42, 45) | 8 | 1(50) | - | 6(29, 57, 60, 64, 66, 69) | 7 | 15 |
| Isolation precautions and their effectiveness for suspected / confirmed patients | 5(14, 23, 26, 27, 88) | - | 1(41) | 6 | 2(47, 77) | 1(82) | 6(57-59, 61, 66, 89) | 9 | 15 |
| Role of personal protective equipment | 1(23) | 1(72) | 1(37) | 3 | 4(46, 47, 76, 77) | 2(53, 83) | 4(55, 56, 58, 60) | 10 | 13 |
| Specific dietary considerations to avoid infection e.g. consumption of wild animals in COVID 19 | 6(13, 23, 26-28, 70) | - | 1(37) | 7 | 1(50) | - | 1(37) | 2 | 9 |
| Role of social distancing measures and preventing mass gatherings e.g. closure of schools | 6(24-29) | 1(82) | - | 7 | - | - | 1(29) | 1 | 8 |
| Measures to adopt when one is ill | 1(22) | 4(30, 33, 36, 84) | 1(39) | 6 | 1(22) | - | - | 1 | 7 |
| Role of avoiding infected or sick patients | 1(29) | 1(82) | 2(45, 82) | 4 | 1(50) | - | 2(29, 57) | 3 | 7 |
| Measures to take after exposure to direct contact with a suspected case | 4(23, 24, 26, 29) | - | 1(79) | 5 | 1(46) | - | 1(61) | 2 | 7 |
| Role of regular cleaning or disinfecting surfaces | 2(25, 70) | - | - | 2 | 4(47, 50, 77, 82) | - | 1(58) | 5 | 7 |
| Role of complementary alternative medicine for prevention of infection e.g. consumption of herbal soups, garlic | 5(24, 25, 28, 29, 70) | - | 1(44) | 6 | - | - | - | 0 | 6 |
| Precautions required post travel e.g. isolation, declaring travel history | 1(29) | 1(72) | 1(39) | 3 | - | - | 3(55, 56, 60) | 3 | 6 |
| Technique and equipment required for proper hand-washing / hand sanitisation | 1(88) | 1(71) | - | 2 | 1(50) | - | 2(55, 56) | 3 | 5 |
| Definitions of terms used in precautionary measures e.g. “safe distance”, “close contacts”, “transient contacts” | 2(70, 88) | - | 1(41) | 3 | - | - | 2(58, 89) | 2 | 5 |
| Necessity for children and young adults to take extra precautions against infection | 3(23, 26, 27) | - | - | 3 | - | - | - | 0 | 3 |
| Role of nasal saline washes or mouthwash | 2(24, 29) | 1(72) | - | 3 | - | - | - | 0 | 3 |
| Role of not touching face, eye or nose | 1(24) | - | 1(45) | 2 | - | - | - | 0 | 2 |
| Role of minimizing travel to affected countries | - | 2(33, 82) | - | 2 | - | - | - | 0 | 2 |
| Role of opening mail carefully | 2(29, 70) | - | - | 2 | - | - | - | 0 | 2 |
| Role of avoiding animals or insects e.g. live animals, mosquitoes | 1(28) | - | 1(37) | 2 | - | - | - | 0 | 2 |
| Role of minimizing infection during meals e.g. use of serving spoons | - | 1(71) | - | 1 | - | - | 1(64) | 1 | 2 |
| Period of self-isolation required for close contacts with known patients | 1(88) | - | - | 1 | - | - | - | 0 | 1 |
| Role of education in preventing spread | - | 1(34) | - | 1 | - | - | - | 0 | 1 |
| Technique and type of face mask to use | 1(88) | - | - | 1 | - | - | - | 0 | 1 |
| Role of healthy lifestyles e.g. exercise | 1(70) | - | - | 1 | - | - | - | 0 | 1 |
| Role of other vaccine | 1(70) | - | - | 1 | - | - | - | 0 | 1 |
| Types of personal protective equipment required for families of suspected / known patients | - | - | - | 0 | - | 1(53) | - | 1 | 1 |
| Role of contact tracing | - | - | - | 0 | - | - | 2(61, 63) | 1 | 1 |
| **Policies and measures implemented by authorities or healthcare institution** |  |  |  |  |  |  |  |  |  |
| Control measures implemented by government e.g. restriction of travel to China | 1(28) | 1(33) | 2(37, 86) | 4 | - | - | 2(61, 63) | 2 | 6 |
| Repercussions associated with non-compliance with measures implemented by government or authorise | 1(28) | - | - | 1 | - | - | - | 0 | 1 |
| Control measures implemented by healthcare institutions | - | - | - | 0 | - | 1(31) | - | 1 | 1 |

**Table 3.** Themes identified from questions related to pandemic attitudes across instruments

| Themes | Number of studies (non-medical populations) |  |  |  | Number of studies (Medical related populations) |  |  |  | Total number |
| --- | --- | --- | --- | --- | --- | --- | --- | --- | --- |
|  | COVID-19 | SARS | MERS | Sub-total | COVID-19 | SARS | MERS | Sub-total |  |
| **Attitudes** |  |  |  |  |  |  |  |  |  |
| *Related to infection in general* |  |  |  |  |  |  |  |  |  |
| Worry, fear, helplessness or anxiety about contracting infection | 4(13, 14, 22, 28) | 4(31, 33, 72, 84) | 1(81) | 10 | 6(22, 49, 51, 52, 76, 77) | 2(31, 83) | 6(60, 61, 63, 65, 68, 78) | 14 | 24 |
| Perceived likelihood of self or others contracting infection | 3(13, 28, 70) | 2(35, 73) | 1(41) | 6 | - | - | 3(52, 60, 82) | 3 | 9 |
| Perceived ability to protect oneself, family members and/or other people around. | 2(28, 70) | 1(30) | - | 3 | 1(87) | - | 1(61) | 2 | 5 |
| Perceived ability in understanding and protecting self and others against the disease outbreak | 3(13, 28, 70) | - | - | 2 | 2(76, 87) | - | - | 2 | 4 |
| Perceived level of self-preparedness for infection outbreak | 2(13, 28) | - | - | 2 | - | - | - | 0 | 2 |
| Perceived impact on daily life | 1(13) |  |  | 1 |  |  | 1(65) | 1 | 2 |
| Belief that infection is preventable | - | - | - | 0 | - | - | 2(61, 63) | 2 | 2 |
| Belief that infection is treatable at home | - | - | - | 0 | - | - | 2(61, 63) | 2 | 2 |
| Beliefs in superstitions, luck or fate | 1(28) | - | - | 1 | - | - | - | 0 | 1 |
| Belief that outbreak will worsen | 1(70) | - | - | 1 | - | - | - | 0 | 1 |
| Belief that discrimination against country of origin is reasonable | 1(70) | - | - | 1 | - | - | - | 0 | 1 |
| Belief that infection is a biochemical weapon developed by foreign countries or terrorists | 1(29) | - | - | 1 | - | - | - | 0 | 1 |
| Feelings of fatigue after outbreak | - | - | - | 0 | 1(77) | - | - | 1 | 1 |
| Acceptance of risk | - | - | - | 0 | - | 1(83) | - | 1 | 1 |
| Avoidance of patients (healthcare workers) | - | - | - | 0 | - | 1(83) | - | 1 | 1 |
| **Related to practices / precautions to minimize transmission of infection** |  |  |  |  |  |  |  |  |  |
| Perceived effectiveness of hospital infection control program in preventing spread | - | - | 1(44) | 1 | 2(48, 52) | - | 6(16, 55, 56, 67, 68, 78) | 8 | 9 |
| Perceived effectiveness of personal protective equipment within healthcare settings | - | - | 1(44) | 1 | 1(48) | - | 6(16, 55, 56, 61, 63, 67) | 7 | 8 |
| Belief that all infected patients should be kept in isolation | 1(14) | - | 1(44) | 2 | 2(48, 52) | - | 4(16, 55, 56, 67) | 6 | 8 |
| Perceived need for intensive care for suspected cases | - | - | - | 0 | 1(48) | - | 5(16, 55, 56, 67, 68) | 6 | 6 |
| Perceived effectiveness of social distancing measure | 2(14, 88) | - | 2(38, 44) | 4 | - | - | - | 0 | 4 |
| Perceived safety of traveling during epidemic/pandemic | 3(14, 28, 70) | - | 1(38) | 4 | - | - | - | 0 | 4 |
| Perceived effectiveness of hand hygiene and good personal hygiene | 2(14, 88) | - | 1(44) | 3 | 1(52) | - | - | 1 | 4 |
| Perceived likelihood of getting vaccination against infection, if available | - | - | - | 0 | 1(52) | - | 3(65, 68, 78) | 4 | 4 |
| Perceived role of health education in disease prevention | - | - | - | 0 | 1(76) | - | 3(29, 61, 63) | 4 | 4 |
| Likelihood of quarantining oneself in infection when symptomatic | 1(14) | - | 1(41) | 2 | - | - | - | 0 | 2 |
| Perceived effectiveness of avoiding infected persons | - | - | 1(44) | 1 | - | 1(83) | - | 1 | 2 |
| Importance of reporting suspected case to health | - | - | - | 0 | - | - | 2(61, 63) | 2 | 2 |
| authorities |  |  |  |  |  |  |  |  |  |
| Perceived effectiveness of avoiding handshaking behaviour | 1(88) | - | - | 1 | - | - | - | 0 | 1 |
| Perceived effectiveness of wearing face mask | - | - | 1(44) | 1 | - | - | - | 0 | 1 |
| Perceived safety of sharing food or eating with other people | 1(88) | - | - | 1 | - | - | - | 0 | 1 |
| Perceived uptake of mask utilization within community | 1(24) | - | - | 1 | - | - | - | 0 | 1 |
| Likelihood of adhering to measures implemented by government or authorities | 1(82) | - | - | 1 | - | - | - | 0 | 1 |
| Perceived level of knowledge on what to do if infected | 1(28) | - | - | 1 | - | - | - | 1 | 1 |
| Perceived likelihood of removing child from school | - | - | 1(41) | 1 | - | - | - | 0 | 1 |
| Avoiding specific dietary intake due to infection (e.g. consumption of non-vegetarian diet) | - | - | - | 0 | 1(76) | - | - | 0 | 1 |
| **Related to healthcare institutions, media, authorities government and universities** |  |  |  |  |  |  |  |  |  |
| Confidence/satisfaction in government or authorities' ability to manage and control of disease outbreak | 4(13, 23, 28, 70) | 3(30, 33, 82) | 1(41) | 8 | 1(77) | - | 5(61, 63, 65, 68, 78) | 6 | 14 |
| Confidence or adequacy in information provided by media, government, authorities or schools about epidemic / pandemic | 3(28, 29, 70) | 2(30, 84) | 1(44) | 6 | - | - | 4(65, 66, 68, 78) | 4 | 10 |
| Perceived need for healthcare workers to be keep up-to-date about pandemic | - | - | 1(44) | 1 | 1(48) | - | 5(16, 55, 56, 68, 78) | 6 | 7 |
| Belief that related information about pandemic should be disseminated to healthcare workers and public | - | - | 1(44) | 1 | 1(48) | - | 4(16, 55, 56, 67) | 5 | 6 |
| Perceived level of understanding on measures adopted by government or authorities and their effectiveness in controlling spread of infection | 1(28) | 1(30) | - | 2 | - | 1(83) | 2(68, 78) | 3 | 5 |
| Belief that government should implement additional measures if cases increases e.g. closure of schools or reduce number of arrivals to Hajj | - | - | 1(38) | 1 | - | - | 2(61, 63) | 2 | 3 |
| Confidence in doctors in accurate diagnosis of infection | - | 1(73) | - | 1 | - | - | - | 0 | 1 |
| Attitudes towards disclosure of exposure to infection by patients | - | - | - | 0 | 1(77) | - | - | 1 | 1 |
| **Impact of pandemic** |  |  |  |  |  |  |  |  |  |
| Perceived severity of infection as a public health threat | 2(13, 70) | 1(72) | 4(39, 41, 44, 45) | 7 | - | - | 3(29, 59, 65) | 3 | 10 |
| Perceived impact of infection on self and/or community | 1(70) | 2(33, 73) | 1(41) | 4 | - | - | 1(61) | 1 | 5 |
| Perceived severe impact of infection on economy | - | 1(33) | - | 1 | - | - | 2(61, 63) | 2 | 3 |
| Perceived risk of job change | - | - | - | 0 | - | 1(83) | - | 1 | 1 |

**Table 4.**
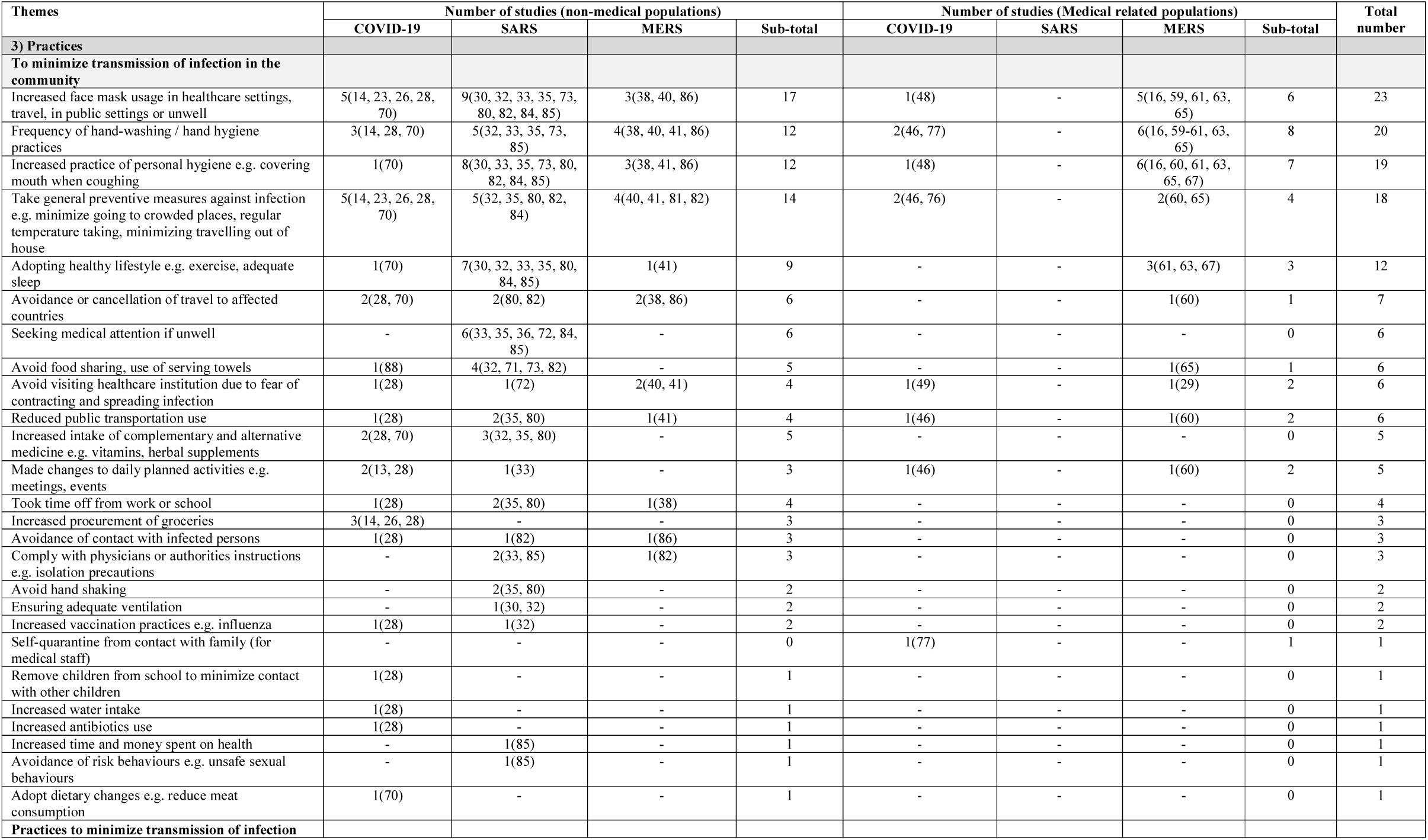
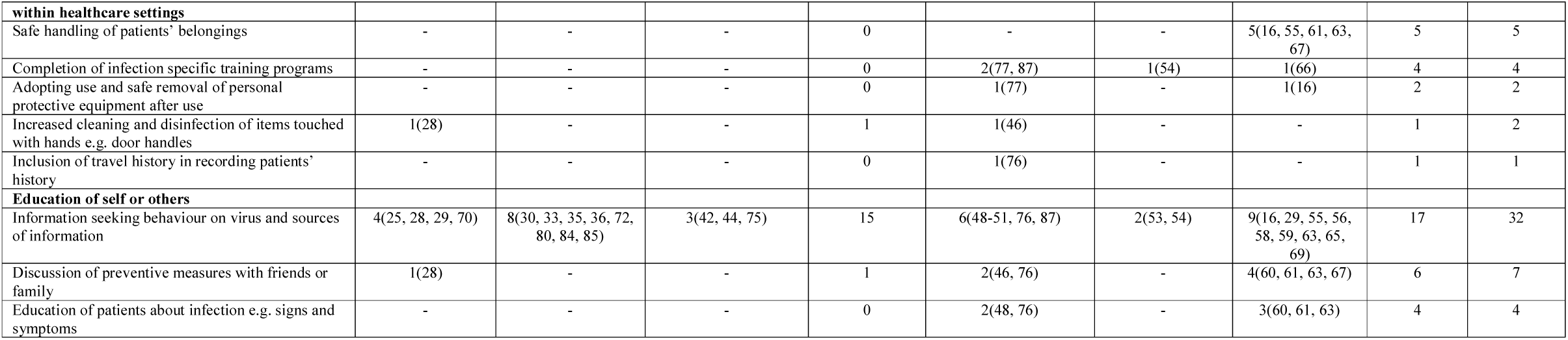
Themes identified from questions related to pandemic practices across instruments

| Themes | Number of studies (non-medical populations) |  |  |  | Number of studies (Medical related populations) |  |  |  | Total number |
| --- | --- | --- | --- | --- | --- | --- | --- | --- | --- |
|  | COVID-19 | SARS | MERS | Sub-total | COVID-19 | SARS | MERS | Sub-total |  |
| **3) Practices** |  |  |  |  |  |  |  |  |  |
| **To minimize transmission of infection in the community** |  |  |  |  |  |  |  |  |  |
| Increased face mask usage in healthcare settings, travel, in public settings or unwell | 5(14, 23, 26, 28, 70) | 9(30, 32, 33, 35, 73, 80, 82, 84, 85) | 3(38, 40, 86) | 17 | 1(48) | - | 5(16, 59, 61, 63, 65) | 6 | 23 |
| Frequency of hand-washing / hand hygiene practices | 3(14, 28, 70) | 5(32, 33, 35, 73, 85) | 4(38, 40, 41, 86) | 12 | 2(46, 77) | - | 6(16, 59-61, 63, 65) | 8 | 20 |
| Increased practice of personal hygiene e.g. covering mouth when coughing | 1(70) | 8(30, 33, 35, 73, 80, 82, 84, 85) | 3(38, 41, 86) | 12 | 1(48) | - | 6(16, 60, 61, 63, 65, 67) | 7 | 19 |
| Take general preventive measures against infection e.g. minimize going to crowded places, regular temperature taking, minimizing travelling out of house | 5(14, 23, 26, 28, 70) | 5(32, 35, 80, 82, 84) | 4(40, 41, 81, 82) | 14 | 2(46, 76) | - | 2(60, 65) | 4 | 18 |
| Adopting healthy lifestyle e.g. exercise, adequate sleep | 1(70) | 7(30, 32, 33, 35, 80, 84, 85) | 1(41) | 9 | - | - | 3(61, 63, 67) | 3 | 12 |
| Avoidance or cancellation of travel to affected countries | 2(28, 70) | 2(80, 82) | 2(38, 86) | 6 | - | - | 1(60) | 1 | 7 |
| Seeking medical attention if unwell | - | 6(33, 35, 36, 72, 84, 85) | - | 6 | - | - | - | 0 | 6 |
| Avoid food sharing, use of serving towels | 1(88) | 4(32, 71, 73, 82) | - | 5 | - | - | 1(65) | 1 | 6 |
| Avoid visiting healthcare institution due to fear of contracting and spreading infection | 1(28) | 1(72) | 2(40, 41) | 4 | 1(49) | - | 1(29) | 2 | 6 |
| Reduced public transportation use | 1(28) | 2(35, 80) | 1(41) | 4 | 1(46) | - | 1(60) | 2 | 6 |
| Increased intake of complementary and alternative medicine e.g. vitamins, herbal supplements | 2(28, 70) | 3(32, 35, 80) | - | 5 | - | - | - | 0 | 5 |
| Made changes to daily planned activities e.g. meetings, events | 2(13, 28) | 1(33) | - | 3 | 1(46) | - | 1(60) | 2 | 5 |
| Took time off from work or school | 1(28) | 2(35, 80) | 1(38) | 4 | - | - | - | 0 | 4 |
| Increased procurement of groceries | 3(14, 26, 28) | - | - | 3 | - | - | - | 0 | 3 |
| Avoidance of contact with infected persons | 1(28) | 1(82) | 1(86) | 3 | - | - | - | 0 | 3 |
| Comply with physicians or authorities instructions e.g. isolation precautions | - | 2(33, 85) | 1(82) | 3 | - | - | - | 0 | 3 |
| Avoid hand shaking | - | 2(35, 80) | - | 2 | - | - | - | 0 | 2 |
| Ensuring adequate ventilation | - | 1(30, 32) | - | 2 | - | - | - | 0 | 2 |
| Increased vaccination practices e.g. influenza | 1(28) | 1(32) | - | 2 | - | - | - | 0 | 2 |
| Self-quarantine from contact with family (for medical staff) | - | - | - | 0 | 1(77) | - | - | 1 | 1 |
| Remove children from school to minimize contact with other children | 1(28) | - | - | 1 | - | - | - | 0 | 1 |
| Increased water intake | 1(28) | - | - | 1 | - | - | - | 0 | 1 |
| Increased antibiotics use | 1(28) | - | - | 1 | - | - | - | 0 | 1 |
| Increased time and money spent on health | - | 1(85) | - | 1 | - | - | - | 0 | 1 |
| Avoidance of risk behaviours e.g. unsafe sexual behaviours | - | 1(85) | - | 1 | - | - | - | 0 | 1 |
| Adopt dietary changes e.g. reduce meat consumption | 1(70) | - | - | 1 | - | - | - | 0 | 1 |
| **Practices to minimize transmission of infection** |  |  |  |  |  |  |  |  |  |
| **within healthcare settings** |  |  |  |  |  |  |  |  |  |
| Safe handling of patients' belongings | - | - | - | 0 | - | - | 5(16, 55, 61, 63, 67) | 5 | 5 |
| Completion of infection specific training programs | - | - | - | 0 | 2(77, 87) | 1(54) | 1(66) | 4 | 4 |
| Adopting use and safe removal of personal protective equipment after use | - | - | - | 0 | 1(77) | - | 1(16) | 2 | 2 |
| Increased cleaning and disinfection of items touched with hands e.g. door handles | 1(28) | - | - | 1 | 1(46) | - | - | 1 | 2 |
| Inclusion of travel history in recording patients' history | - | - | - | 0 | 1(76) | - | - | 1 | 1 |
| **Education of self or others** |  |  |  |  |  |  |  |  |  |
| Information seeking behaviour on virus and sources of information | 4(25, 28, 29, 70) | 8(30, 33, 35, 36, 72, 80, 84, 85) | 3(42, 44, 75) | 15 | 6(48-51, 76, 87) | 2(53, 54) | 9(16, 29, 55, 56, 58, 59, 63, 65, 69) | 17 | 32 |
| Discussion of preventive measures with friends or family | 1(28) | - | - | 1 | 2(46, 76) | - | 4(60, 61, 63, 67) | 6 | 7 |
| Education of patients about infection e.g. signs and symptoms | - | - | - | 0 | 2(48, 76) | - | 3(60, 61, 63) | 4 | 4 |

For attitudes about pandemics, worry/fear/helplessness about pandemic (13, 14, 22, 28, 31, 33, 49, 51, 52, 60, 61, 63, 65, 68, 72, 76-78, 81, 83, 84), confidence in governments’ ability to manage pandemic (13, 23, 28, 30, 33, 41, 61, 63, 65, 68, 70, 77, 78, 82) and perceived severity of infection as a public health problem (13, 29, 39, 41, 44, 45, 59, 65, 70, 72) was most commonly assessed. (Table 3)

For practices in pandemics, behaviours related to mask utilization (14, 16, 23, 26, 28, 30, 32, 33, 35, 38, 40, 48, 59, 61, 63, 65, 70, 73, 80, 82, 84-86), hand hygiene (14, 16, 28, 32, 33, 35, 38, 40, 41, 46, 59-61, 63, 65, 70, 73, 77, 85, 86), personal hygiene (16, 30, 33, 35, 38, 41, 48, 60, 61, 63, 65, 67, 70, 73, 80, 82, 84-86) and information seeking (16, 25, 28-30, 33, 35, 36, 42, 44, 48-51, 53-56, 58, 59, 63, 65, 69, 70, 72, 75, 76, 80, 84, 85, 87) were most commonly studied. (Table 4)

Figure 2 shows the proposed framework (PANDEMIC-HL) for items to be included in generic pandemic health literacy tools.

**Figure 2.**
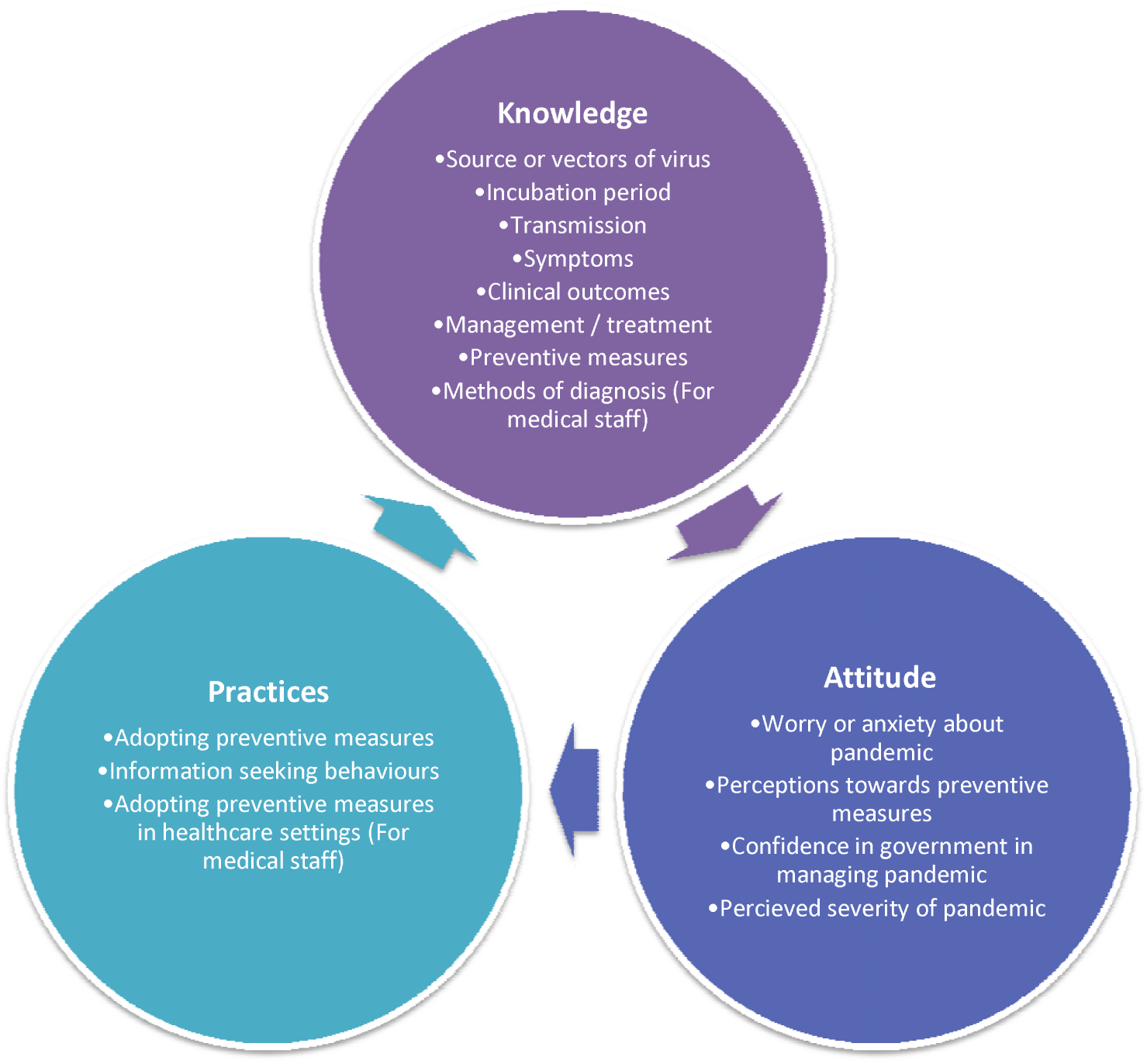
Proposed framework for items included in PANDEMIC related Health Literacy instruments (PANDEMIC-HL)

### Determinants of health literacy

Across the five domains for health literacy related factors, 34 factors were identified. (Table 5) Among these, socio-demographic-economic and health systems-based domains were the most studied. Socio-demographic factors which were commonly associated with better health literacy included higher educational level (23, 26, 27, 33, 34, 36, 37, 39, 40, 42, 43, 45, 72-77, 80-82, 88, 89), increased age (23, 26, 30, 37, 40, 43, 48, 50, 63, 65, 72, 74, 75, 78, 84, 85) and female gender (13, 16, 23, 26, 33, 34, 38, 40, 41, 44, 45, 60, 62, 72, 74, 78, 80, 81, 84, 85, 89). For health systems-based factors, increased experience in the healthcare system (48, 56, 63, 76, 77, 89) and attendance in health education programs (28, 33, 58, 60, 71, 78) were associated with better health literacy. For medical and psychiatric/psychological factors, increased general health literacy (13, 30) and increased anxiety about the spread of infection (28, 33, 80, 84) were associated with better health literacy. Lastly, other factors associated with better health literacy included the use of traditional sources of information such as newspaper or television (28, 33, 75).

**Table 5.** Factors and their association with better health literacy (Knowledge, Attitudes and Practices)

| Factors | Total number of supporting studies (N, Cited studies) |  |  | Total |
| --- | --- | --- | --- | --- |
|  | COVID-19 | SARS | MERS |  |
| **1) Socio-demographic factors** |  |  |  |  |
| *Socio-demographics* |  |  |  |  |
| Education level |  |  |  | **26** |
| • ↑ Education level | 6(23, 26, 27, 76, 77, 88) | 7 (33, 34, 36, 72, 73, 80, 82) | 10 (37, 39, 40, 42, 43, 45, 74, 75, 81, 89) | 23 |
| • ↓ Education level | 1(26) | 2 (72, 80) | - | 3 |
| Age |  |  |  | **26** |
| • ↑ Age | 4(23, 26, 48, 50) | 4(30, 72, 84, 85) | 8(37, 40, 43, 63, 65, 74, 75, 78) | 16 |
| • ↓ Age | 1(48) | 4(32, 33, 36, 82) | 5(37, 41, 42, 44, 81) | 10 |
| Gender |  |  |  | **25** |
| • Female | 3(13, 23, 26) | 6(33, 34, 72, 80, 84, 85) | 12(16, 38, 40, 41, 44, 45, 60, 62, 74, 78, 81, 89) | 21 |
| • Male | 1(46) | - | 3(16, 55, 56) | 4 |
| Marital status (Married vs unmarried, divorced) | 3(13, 23, 27) | 2(34, 85) | 2(40, 74) | **7** |
| Race (e.g. White vs Black, Chinese and Malay vs Others) | 1(26) | 3(33, 36, 84) | 1(75) | **5** |
| Discipline of study (Medical vs non-medical) | 1(50) | - | 4(29, 37, 44, 66) | **5** |
| Nationality (Finland vs Dutch, Korean, Jordan vs Saudi Arabian) | - | 1(35) | 4(41, 58, 79, 81) | **5** |
| Pilgrims (vs non-pilgrims) | - | - | 2(39, 79) | **2** |
| Type of residence (Villa vs flat, Apartments with more rooms vs less rooms) |  | 2 (34, 36) | 1(75) | **3** |
| Level of English proficiency | 1(13) | - | - | **1** |
| Location of residence (Hubei vs Other parts of China) | 1(23) | - | - | **1** |
| *Economic / employment related* |  |  |  |  |
| Employed (vs unemployed) | 4(13, 23, 27, 88) | 2 (34, 36) | 2 (42, 74) | **8** |
| Healthcare workers (vs non healthcare workers) | 4(48, 50, 52, 77) | - | 3(43, 63, 78) | **7** |
| Income level |  |  |  | **6** |
| • ↑ Income level | 2(26, 88) | 1 (34) | 1 (40) | 4 |
| • ↓ Income level | 2(26, 27) | - | - | 2 |
| Type of employment (Mental labour vs unemployed, physical labour, students) | 1(23) | - | - | **1** |
| **2) Medical factors** |  |  |  |  |
| ↑ General health literacy | 1(13) | 1(30) | - | **2** |
| Self-reported health status (good to excellent vs poor) | 1(13) | - | - | **1** |
| ↓ Number of chronic conditions | 1(13) | - | - | **1** |
| ↑ Health activation (an individual's willingness to take on the role of managing their health and healthcare) | 1(13) | - | - | **1** |
| ↑ Self-efficacy in performing preventive health practices | - | 1(30) | - | **1** |
| **3) Psychological / psychiatric factors** |  |  |  |  |
| ↑ Anxiety related to spread of infection | 1(28) | 3(33, 80, 84) | - | **4** |
| ↑ Perceived susceptibility to being infected | - | 1(30) | 1(60) | **2** |
| ↓ Superstition and fatalism | 1(28) | - | - | **1** |
| **4) Health systems-based factors** |  | - | - |  |
| Years of experience in healthcare system |  |  |  | **7** |
| • ↑ Years of experience in healthcare system | 3(48, 76, 77) | - | 3(56, 63, 89) | **6** |
| • ↓ Years of experience in healthcare system | 1(46) | - | - | **1** |
| Attendance in health education programs (Public health prevention programs, continuous medical education activities) | 1(28) | 2(33, 71) | 3(58, 60, 78) | **6** |
| **Physicians' specialty or place of practice** |  |  |  | **3** |
| Non-emergency department (vs emergency Department) | 1(46) | - | - | **1** |
| Private Medical Sector (vs Public Medical Sector) | - | - | 1(89) | **1** |
| Specialist (vs Primary Care Physicians) | - | - | 1(58) | **1** |
| Presence of infection control programmes in hospitals | - | - | 1(78) | **1** |
| **5) Others** |  |  |  |  |
| **Sources of information** |  |  |  |  |
| Traditional Media (TV, Newspaper Radio Sources) vs others | 1(28) | 1(33) | 1(75) | **3** |
| Social Media vs others | 1(28) | - | - | **1** |
| Textbook and lectures vs others | - | - | 1(58) | **1** |
| **Political or government related factors** |  |  |  |  |
| General confidence in government / authority |  |  |  | **2** |
| • ↑ General confidence in authority | - | 1(33) | - | **1** |
| • ↓ General confidence in authority | 1(28) | - | - | **1** |
| Political affiliation (Democrats and independents vs republicans) | 1(26) | - | - | **1** |

### Clinical outcomes

Among the included studies, no studies evaluated clinical outcomes related to COVID-19, SARS or MERS.

## Discussion

This review provided a summary of existing literature regarding health literacy regarding COVID-19, SARS and MERS as well as factors and clinical outcomes associated with poor health literacy. To our best knowledge, this is the first review to summarise health literacy in the COVID-19, SARS and MERS pandemics.

Overall, the level of health literacy related to COVID-19 and other pandemics remains sub-optimal in both medical and non-medical populations. Given the important role health literacy plays in stemming the spread of infection and mitigating the impacts of these pandemics, there is an urgent need for the design of interventions to rapidly enhance the pandemic related health literacy of the population. Within the field of health literacy for non-communicable diseases, both single and mixed strategies encompassing interventions such as alternative readability and document design, alternative numerical presentation, pictorial representation and use of alternative media have been employed.(90) Alternative document design has been adapted in the construction of health information websites where the use of simple designs, minimization of lengthy text and medical jargon use have been shown to enhance health literacy among users.(91) These interventions are relevant and should be adapted for pandemic related health literacy.(90) In the current information technology era, websites, web-based applications and mobile applications serve important vehicles for the dissemination of critical pandemic related information. A review by Kim et al. highlighted readability and other resource-specific factors e.g. accessibility, interactivity and comprehensiveness as barriers to online health information users.(92) Of note, the readability of most online health resources exceeded the recommended sixth-grade reading level.(92) The readability of online health resources related to the COVID-19 has not been evaluated, and future researchers should consider utilizing instruments such as Simplified Measure of Gobbledygook or Flesch Reading Ease for the evaluation of these resources.(93, 94)

For populations at increased risk of poor clinical outcomes of infections such as the elderly, immunocompromised patients, human-immunodeficiency virus or with multiple comorbidities, they form high priority populations where the levels of pandemic health literacy should be assessed.(95, 96) In the context of the current COVID-19 pandemic, our review only found 1 study which specifically evaluated the health literacy related to COVID-19 among these high-risk populations.(13) It is imperative that future research is undertaken to evaluate the health literacy among these patient populations for targeted interventions to be designed for patients if required.

Significant heterogeneity in the instruments used for the assessment of pandemic related health literacy was noted in our review. Currently, there is no gold-standard instrument which evaluates pandemic-related health literacy. Given the need for validated and standardised tools to be created to facilitate the evaluation of pandemic related health literacy, the PANDEMIC-HL framework was proposed to guide the selection of topics to be addressed in instruments. It was modelled after psychosocial models of health behaviours and encompassed key topics which were frequently evaluated in instruments across the three pandemics.(97) It is hoped that the framework will serve as a foundation for facilitating the development of health literacy tools for future pandemics.

A detailed evaluation revealed critical issues faced in the design of COVID-19 related health literacy questionnaires, especially in the wake of evolving information around the new pandemic. For example, one of the questions in the survey used by Moro et al. described the practice of not travelling to China as the only correct answer for preventing oneself from contracting COVID-19, amidst potentially correct answers such as avoiding crowded places. While this was understandable as the study was conducted in the early stages of COVID-19 outbreak prior to the implementation of social distancing measures, it showcases the importance of being up-to-date with the latest pandemic related information for researchers designing future health literacy trials. Studies that are ongoing within the midst of pandemics should also check regularly that the items within questionnaires and their answers reflect the current state of the evidence, as emerging new information may lead to inaccuracies in the assessment of health literacy. With the implementation of lockdowns in countries to prevent the transmission of COVID-19, the use of online surveys and questionnaires has been increasingly used for pandemic related health literacy research. The results of these studies should be evaluated carefully given the following limitations. Firstly, as the recruited participants are limited to those who are keyboard literate, this may limit the generalizability of the study results.(98) Additionally, the validity of these study results may be affected as these surveys commonly suffer from poor response rates.(98)

With regards to the determinants of pandemic related health-literacy, higher education levels, older age, female gender, and being employed were the most studied factors associated with higher pandemic related health literacy. Our results generally concurred with the determinants of general health literacy based on current literature. Pertaining to the role of gender, females have identified in multiple studies to have higher general health literacy levels as compared to their male counterparts. (99, 100) This difference may be related to the traditional roles that females play in caring for family members and children, which increase their need and familiarity with navigating and interacting with healthcare information and systems. (101) Employment creates opportunities for individuals to access healthcare resources..(17) Likewise, the education attainment plays an important role in health literacy through its influence on knowledge, skills and resource interpretation and utilization.(10, 102) Interestingly, while older age has been associated with poorer health literacy in the general population possibly due to ageing-related factors such as cognitive decline and physical impairments(103), our review showed that older age was associated with better health literacy. This may be related to the greater number of pandemics an older person experience in his lifetime where prior knowledge gained from previous pandemics may shape their ability to gather, synthesize and comprehend information related to ongoing pandemics. (10, 104) Another potential reason for this finding could be related to increased selection bias among older participants as compared to studies performed for general health literacy. While our review has highlighted multiple determinants of pandemic related health literacy, more studies are required to understand the complex interplay between these factors and their impact on health literacy.

In our review, there were no studies which evaluated clinical outcomes associated with poor pandemic related health literacy. Poor general health literacy has been linked to adverse clinical outcomes such as increased healthcare utilization and morbidity.(105) In addition, people with low general health literacy are more likely to delay or forego medical treatment, compared to their counterparts with adequate health literacy. (106) While it is expected that people with poor pandemic related health literacy may have poorer clinical outcomes, this remains a significant research gap that should be addressed in future studies.

The main strength of this review was that health literacy in previous coronaviruses related pandemics such as SARS and MERS were evaluated to provide a more comprehensive overview of pandemic related health literacy. However, the findings from this review should also be interpreted with the following limitations. Firstly, while we adopted a reasonably comprehensive search strategy, potentially relevant articles may have been missed. Finger searching within the references of included articles was performed to minimize this omission of potentially relevant articles. Secondly, we were only able to include articles in the English language due to the language limitations of the authors. Thirdly, we were not able to perform meta-analyses for the overall level of pandemic related health literacy and their determinants due to the heterogeneity in instruments. With the development of a standardised instrument for the assessment of health literacy related to pandemics, future studies should consider using meta-analyses to compare the level of health literacy across different populations. Lastly, the full questionnaires could not be accessed for 27 studies. While themes described in the main text of these articles were carefully extracted, we could not rule out the omission of themes which were not described. Future health literacy studies should append their questionnaires to allow meaningful evaluation of the study results.

## Conclusion

Overall, the level of pandemic related health literacy remains sub-optimal among both the medical and non-medical population. This is worrisome given the critical role health literacy serves in reducing the spread of contagion and mitigating the effects of pandemics. There is an urgent need to develop up-to-date, validated and standardised questionnaires for the rapid assessment of pandemic-related health literacy. Important determinants associated with better levels of health literacy such as older age, female gender, employment status and education level were highlighted in this review. Healthcare administrators and policymakers need to be mindful of these determinants when formulating dissemination of critical pandemic related information and interventions to improve the health literacy of the population. More studies are required to evaluate the clinical outcomes associated with pandemic related health literacy.

## Data Availability

The data referred to in the manuscript are available from the results, tables and supplementary files.

## Authors’ Contributions

JJB Seng was the study’s principal investigator and was responsible for the conception, initial literature review and design of the study. CT Yeam, CW Huang, NC Tan and LL Low were the co-investigators. JJB Seng, CT Yeam and CW Huang were responsible for the screening and inclusion of articles and data extraction. All authors contributed to the data analyses and interpretation of data. JJB Seng prepared the initial draft of the manuscript. All authors revised the draft critically for important intellectual content and agreed to the final submission.

## Acknowledgements

None

## Guarantor’s name

JJB Seng is the guarantor of this work and, as such, had full access to all the data in the study and takes responsibility for the integrity of the data and the accuracy of the data analysis.

## Funding and role of funding source

This research did not receive any specific grant from funding agencies in the public, commercial, or not-for-profit sectors.

## Conflict of interest

The authors declare that we have no conflict of interests.

## Ethics approval

This study was exempted from institutional ethics board approval as it is a systematic review of existing literature.

## Availability of data and materials

The data and materials used in the study have been presented in this review.

## Notes

### Competing Interest Statement

The authors have declared no competing interest.

### Clinical Trial

nil

### Funding Statement

This research did not receive any external funding

